# Magnetic resonance imaging for individual prediction of treatment response in major depressive disorder: a systematic review and meta-analysis

**DOI:** 10.1101/2020.06.27.20141465

**Authors:** Sem E. Cohen, Jasper B. Zantvoord, Babet N. Wezenberg, Claudi L.H. Bockting, Guido A. van Wingen

## Abstract

**Objective:** No tools are currently available to predict whether a patient suffering from major depressive disorder (MDD) will respond to a certain treatment. Machine learning analysis of magnetic resonance imaging (MRI) data has shown potential in predicting response for individual patients, which may enable personalized treatment decisions and increase treatment efficacy. Here, we evaluated the accuracy of MRI-guided response prediction in MDD.

**Methods:** We conducted a systematic review and meta-analysis of all studies using MRI to predict single-subject response to antidepressant treatment in patients with MDD. Classification performance was calculated using a bivariate model and expressed as area under the curve, sensitivity, and specificity. In addition, we analyzed differences in classification performance between different interventions and MRI modalities.

**Results:** Meta-analysis of twenty-two samples including 957 patients showed an overall area under the bivariate summary receiver operating curve of 0.84 (95% CI 0.81-0.87), sensitivity of 77% (95% CI 71-82), and specificity of 79% (95% CI 73 - 84). Although classification performance was higher for electroconvulsive therapy outcome prediction (n = 285, 80% sensitivity, 83% specificity) than medication outcome prediction (n = 283, 75% sensitivity, 72% specificity), there was no significant difference in classification performance between treatments or MRI modalities.

**Conclusion:** Prediction of treatment response using machine learning analysis of MRI data is promising but should not yet be implemented into clinical practice. Future studies with more generalizable samples and external validation are needed to establish the potential of MRI to realize individualized patient care in MDD.

**PROSPERO registration number:** CRD42019137497

## Introduction

Major depressive disorder (MDD) is a debilitating disease, accounting for 40 percent of the global disability-adjusted life years caused by psychiatric disorders (1). Depression is associated with impaired social functioning and unemployment, and is a causal factor in a wide range of chronic physical illnesses such as diabetes and cardiovascular disease (2, 3). MDD is estimated to have a life-time prevalence of 20,6 percent in the United States (4). Despite general consensus that effective treatment of depression is paramount for both a patient’s health and for reducing global burden of disease, MDD disease burden has not decreased in the last decades (5). This is partly because treatment selection is based on trial and error, with no possibility to predict an individuals’ response to certain treatment (6). Non-response to initial pharmacological and psychotherapeutic interventions is highly prevalent, with treatment-resistant depression affecting 20-30 percent of depressed patients in current clinical practice (7-9). Treatment of choice for patients who have not responded to pharmacological and psychotherapeutic treatments is electroconvulsive therapy (ECT), which produces remission rates in about 50 percent of therapy resistant patients (10, 11). Furthermore, non-response can only be determined at least four weeks after initiation of pharmacotherapy, ECT requires 4-6 weeks on average and effects of psychotherapy can even take sixteen weeks to manifest (7, 12). Consequently, patients are regularly exposed to multiple unsuccessful treatments and might spend months to years waiting for adequate treatment. This stresses the need for markers which, before treatment commencement, can inform clinicians on the chance of responding to a particular treatment.

A large number of studies have correlated baseline clinical characteristics and biomarkers with MDD status and treatment outcome, and have identified many factors that are associated with treatment success (13) (14). However, such descriptive analyses only provide inference at the group level and not at the level of the individual patient, which is required for clinical decision making (15). More recent studies have started to use machine learning analyses that aim to develop predictive models, and which are tested using independent data (13). More than with correlational analysis, single-subject response prediction studies using machine learning might be able to redeem the promise of individualized psychiatry (16). Without being explicitly pre-programmed, these algorithms are able to learn from aggregated data in a patient sample using multivariate pattern recognition, in order to provide the best prediction of an output variable (17, 18). Machine learning algorithms could enable clinicians to judge the viability of treatments for individual patients. As such it might increase treatment efficacy, decrease illness duration and reduce MDD disease burden.

Multiple modalities have been considered for single-subject response prediction. A recent review covering different markers, found neuroimaging to overall be most successful in predicting treatment response in depressed patients (i.e. more than phenomenological or genetic studies) (19).However, the review pooled different treatments and neuroimaging modalities such as electroencephalography (EEG) and magnetic resonance imaging (MRI), and thus offers little insight into treatment specific biomarkers or specific imaging modalities. A recent meta-analysis on EEG for individual prediction of antidepressant treatment response found reasonable accuracy (72% sensitivity and 68% specificity) but concludes that EEG should not yet be used clinically as a prediction tool, since generalizability and validity of literature is limited (20). However, a meta-analysis of single-subject MRI-studies for prediction of anti-depressive treatment does not yet exist.

The primary aim of the present study was to calculate the aggregate classification performance of predictive MRI biomarkers in patients with MDD using a bivariate random effect model meta-analysis. We further investigated if classification performance was influenced by intervention type (i.e. pharmacotherapy, psychotherapy or ECT) or imaging modality (i.e. structural MRI, resting-state fMRI, task -based fMRI, DTI).

## Methods

### Inclusion and exclusion criteria

Two authors (SC and BW) included studies using any form of MRI (structural, resting-state, task-based, spectroscopy, diffusion tensor imaging) which were conducted at baseline, i.e. within 4 weeks before start of antidepressant treatment. Furthermore, inclusion criteria were an overarching definition of antidepressant treatment according to the current NICE-guidelines and a non-selective patient population with MDD suffering from a current depressive episode. Studies which used feature selection based on in-sample data without validating prediction outcomes either internally (e.g. through cross-validation) or externally (through independent set validation), were excluded. In- or exclusion conflicts were resolved by consensus, or if necessary by authors JZ and GW.

### Search strategy

We conducted a search in EMBASE, Medline, PsycInfo and Web of Science databases. Each database was searched from inception to January 2020. Furthermore, we searched the WHO International Clinical Trial Registry Platforms search portal for registered and unpublished studies and we looked for ‘grey’ literature such as abstracts and conference articles through conference websites and from other relevant sources. Additionally, we checked included articles for references and conducted citation screening. For a full account of our search strategy and inclusion criteria, see the supplementary material.

### Data extraction

Two authors (SC and BW) independently extracted data from included studies, including: number of participants, patient population and depression severity subtype, treatment history, antidepressant intervention and outcome measures, response/remission rates, neuroimaging technique, brain region and feature selection, method of analysis and validation strategy (see table 1). From included articles we extracted the confusion table (a 2×2 table for correctly and incorrectly classified patients) for sensitivity or specificity. If these were not supplied, we computed the matrix from additional information in the article. If multiple studies analyzed the same patient sample, we used mean outcome measures based on these studies. If necessary, we contacted authors requesting additional information.

**Table 1:** methodological summary of studies.

| Study + year | n | outcome | intervention | duration | modality | analysis | validation |
| --- | --- | --- | --- | --- | --- | --- | --- |
| Costafreda, 2009-1 | 16 | remission | CBT | 16 wk | tbfMRI | SVM | LOO CV |
| Siegle, 2012 | 12 | response | CBT | 12 wk | tbfMRI | RF | Ind. replication |
| Queirazza, 2019 | 37 | response | CBT | 6-10 wk | tbfMRI | SVM, LR | LOO nested CV |
| Van Waarde, 2015 | 45 | remission | ECT | 10 wk | rsfMRI | ISVM | LOO CV |
| Moreno-Ortega, 2019 | 19 | remission | ECT | ns | rsfMRI | LR | LOO CV |
| Sun, 2019 | 122 | remission + remission | ECT | 3-4 wk | rsfMRI | LR | LOO CV |
| Redlich, 2016 | 23 | response | ECT | 3-8 wk | sMRI | ISVM/ GPC | LOO CV |
| Wade, 2016 | 34 | response | ECT | 2-7 wk | sMRI | RBFSVM | LOO CV |
| Cao, 2018 | 24 | response + remission | ECT | 3-4 wk | sMRI | ISVM | LOO CV |
| Jiang, 2018 | 38 | remission | ECT | 3-4 wk | sMRI | LR | 10-fold LOO CV + Independent cohort rep |
| Wade, 2017 | 44* | remission | ECT | ns | sMRI | RF | Nested CV |
| Leaver, 2017 |  | response | ECT | ns | rsfMRI, aslMRI | RBFSVM | 5-fold LOO CV |
| Drysdale, 2017 | 124 | response | rTMS | 4-6 wk | rsfMRI | ISVM | LOO CV |
|  | 30 |  |  |  |  |  | Ind. replication |
| Cash, 2019 | 33 | remission | rTMS | 5-8 wk | rsfMRI | ISVM | LOO + k fold CV |
| Costafreda, 2009-2 | 18* | remission | SSRI | 8 wk | sMRI | ISVM | LOO CV |
| Nouretdinov, 2011 |  | remission | SSRI | 8 wk | sMRI | TCP | LOO CV |
| Gong, 2011 | 46 | response | SSRI/TCA/ SNRI | 12 wk | sMRI | ISVM | LOO CV |
| Marquand, 2008 | 20 | response | SSRI | 8 wk | tbfMRI | ISVM | LOO CV |
| Godlewska, 2018 | 32 | response | SSRI | 6 wk | tbfMRI | LR | LOO CV |
| Meyer, 2019 | 22 | remission/ nonresponse | SSRI | 8 wk | tbfMRI | LR | LOO CV |
| Karim, 2018 | 49 | remission | SNRI | 12 wk | tbfMRI | LR | 10-fold LOO CV |
| Patel, 2015 | 19 | remission | SSRI/ SNRI | ns | rsfMRI, DTI, sMRI | ADTree/ ISVM/ RBFSVM/ L1LR | Nested LOO CV |
| *iSPOT trials* | 77* |  | SSRI/SNRI | 8 wk |  |  |  |
| Korgaonkar, 2014 |  | remission |  |  | DTI | LR | K-fold CV |
| Williams, 2015 |  | response |  |  | tbfMRI | LDA | LOO CV |
| Goldstein-Piekarski, 2016 |  | remission |  |  | tbfMRI | LR | 10-fold LOO CV |
| Grieve, 2016 |  | non-remission |  |  | DTI | LR | independent rep |
| Goldstein-Piekarski, 2018 |  | remission |  |  | rsfMRI | LR | LOO CV |

\* n is a weighted average across studies. Reported sample sizes were not necessarily equal in articles with overlapping sample. SSRI = selective serotonin reuptake inhibitor, TCA = tricyclic antidepressant, SNRI = serotonin-norepinephrine reuptake inhibitor, ECT = electroconvulsive therapy, CBT = cognitive behavioral therapy, rTMS = repetitive transcranial magnetic stimulation, iTBS = intermittent theta burst stimulation, AP = antipsychotics, ns = not specified, tb = task-based, rs = resting state, asl = arterial spin labeling, fMRI = functional magnetic resonance imaging, sMRI = structural magnetic resonance imaging, WB = whole-brain, ROI = region-of-interest, DTI = diffuse tensor imaging, ISVM = linear support vector machine, RBF = radial basic function, TCP = Transductive conformal predictor, LR = logistic regression, LinR = linear regression, LDA = linear discriminant analysis, RF = random forest, LOO CV = leave-one-out cross-validation, wm = white matter, sLR = stepwise linear regression, beta-w = beta-weights, LARS = least-angle regression PMVD = proportional marginal decomposition

### Meta-analytic method

For quantitative analysis, we used confusion matrices to pool studies using Reitsma’s bivariate random effect model, as suggested in the Cochrane handbook for diagnostic tests of accuracy studies (21, 22). We used this method for computing our main outcomes, which were the overall area under the summary receiver operating characteristic (ROC) curve, sensitivity, and specificity, as well as sensitivity and specificity of intervention subsets. Additionally, we performed a separate bivariate regression for modalities (functional and structural MRI) by including from each study both sMRI and fMRI, if provided in the original article or after our request for further information.

### Heterogeneity and publication bias

To visualize between-study-differences, we conducted a univariate random-effect forest plot of the diagnostic odds ratios, subdivided per treatment group. We identified clinical and statistical heterogeneity by visually assessing confidence interval overlap and by identifying outlying studies. We avoided using an objective measure of heterogeneity, since these have shown to be inappropriately conservative for accuracy studies (23). Rather, we used a random-effect model that assumes that our data was heterogeneous and set out to investigate potential sources of heterogeneity (22). We did not perform any sensitivity analyses, as no studies were of such low quality, or were such outliers that sensitivity analysis was appropriate. To assess sample size effects and possible publication bias, we used Deeks’ test, as recommended for diagnostic accuracy studies (24, 25). For assessing quality of the primary studies, we used the QUADAS-2 tool (26). We pre-specified methods in the PROSPERO database for systematic reviews (registration number CRD42019137497) (ref naar prospero). All analyses were conducted using the mada and metafor package in R.

## Results

### 3.1 Search results

Our search yielded 5624 hits, 168 of which were included for full-text review (see **figure 1**). After contacting the authors for additional information, we excluded 21 studies for not reporting data necessary for reconstructing a confusion matrix, all of which were ‘grey literature’, i.e. abstracts or conference summary articles. Furthermore, we excluded 11 articles for not reporting any form of validation of their prediction model. After exclusion of non-eligible studies and, through citation searching, addition of two eligible studies which did not come up in search hits, 27 remained (27-53).

**Figure 1:**
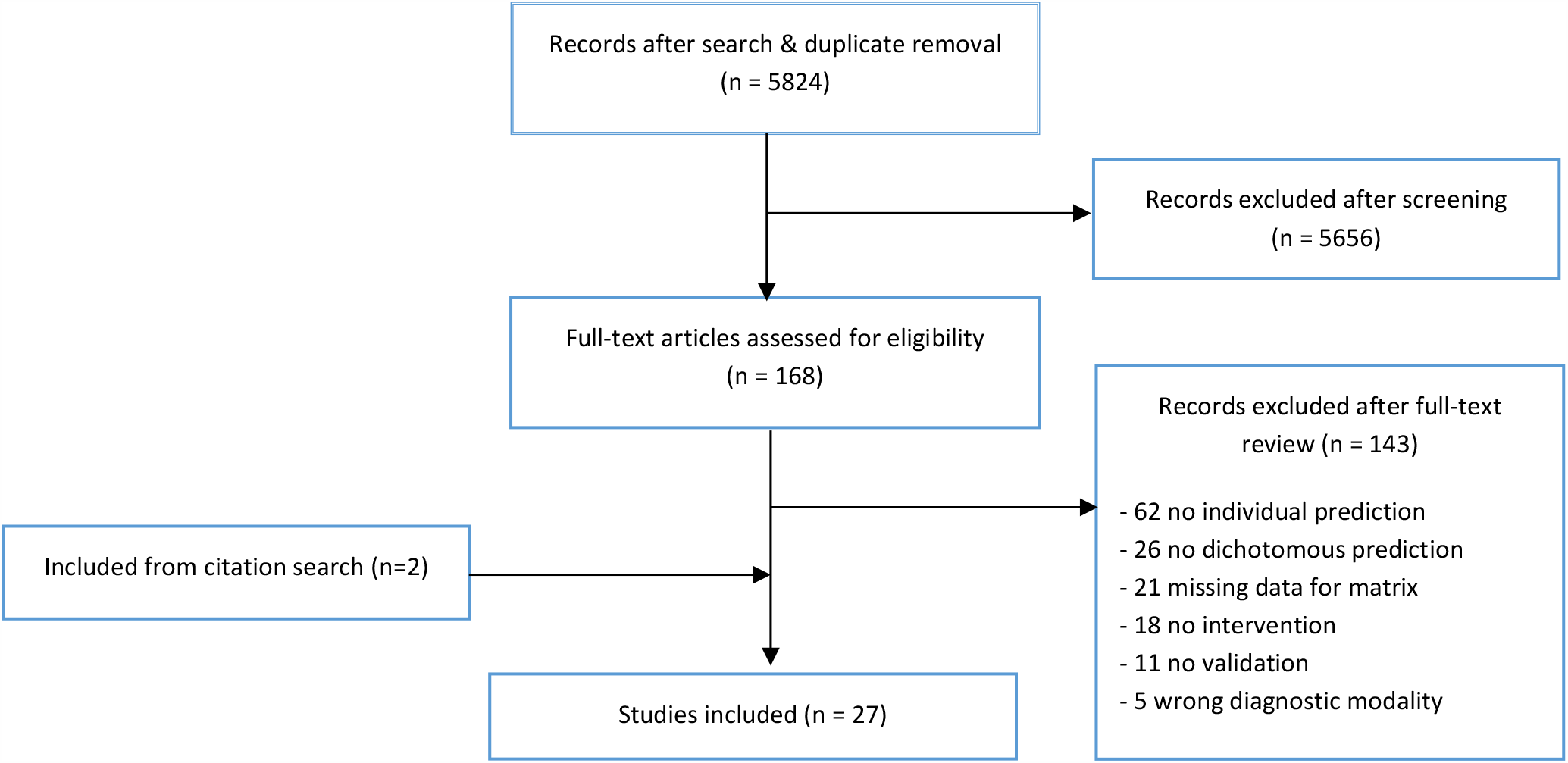
flow-diagram of study inclusion process.

### 3.2 Description of study characteristics

We included 27 studies with an accumulated number of 957 unique patients and a mean sample size of 44 per study, with a median of 33 (see **Table 1** and **Supplementary table 1** for a full methodological study summary). Three patient samples were used in more than one article (27, 29, 37, 38, 48-52).

Of the included studies, 50% used some form of pharmacotherapeutic intervention (total n = 283), all of which administered a clinically viable dosage, with response time varying from two weeks (early response), to 12 weeks. ECT was administered in 35% of studies (total n = 285), 8% used transcranial magnetic stimulation and 8% used cognitive therapy. Most studies used either sMRI (31%) or task-based fMRI (31%), most often using emotional stimuli, 19% used resting-state fMRI and 8% used diffusion tensor imaging (DTI). Two studies combined multiple modalities (37, 47).

As machine learning paradigm, 31% studies used support vector machine (SVM) for data-analysis, while 28% used logistic regression. After comparing classification accuracy with multiple algorithms (among others SVM and random forest) Patel and colleagues used an alternating decision tree method (47). For validation, 85% used leave-one-out cross-validation. Two studies used an independent cohort to validate their results, while one study first cross-validated classification results, after which authors validated their prediction model in two small, independent cohorts, achieving similar results (36, 40, 50). For additional information on approaches to imaging analysis, please refer to **supplementary table 2**.

### 3.3 Meta-analysis

#### 3.3.1 General outcome

After pooling results from studies with overlapping patient samples, we quantitatively analyzed 22 samples, including one independent cohort replication which we have interpreted as a separate study (40). For all imaging modalities and interventions taken together, the meta-analytic estimates for the sROC AUC was 0.84 (95% CI 0.81-0.87), with 77% sensitivity (95% CI 71-82) and 79% specificity (95% CI 73 - 84), amounting to a moderately high classification performance.

#### 3.3.2 Intervention differences

Sensitivity and specificity of ECT-interventions were 80% (95% CI 73-85) and 83% (95% CI 72-90), respectively, compared to 75% (95% CI 68-82) and 72% (95% CI 64-80) for antidepressant medication. Although prediction outcomes in ECT-studies do show a trend towards higher precision, confidence intervals overlap (see **table 2**).

**Table 2:** Summary estimates of sensitivity/specificity for different interventions.

| Intervention group | Sensitivity | 95% CI | Specificity | 95% CI |
| --- | --- | --- | --- | --- |
| Combined | 77% | 71-82 | 79% | 73 - 84 |
| Medication | 75% | 68- 82 | 73% | 64 - 80 |
| ECT | 80% | 73 – 85 | 83% | 72 - 90 |
| Psychotherapy | 84% | 68 - 92 | 72% | 39 - 92 |
| rTMS | 79% | 71 – 86 | 82% | 74 - 88 |

CI = confidence interval, rTMS = repetitive
transcranial magnetic stimulation

#### 3.3.3 Modality differences

In order to assess whether sMRI studies yielded different performance measures compared to fMRI studies, we performed random-effect meta-regression for modality subtypes. When comparing fMRI and sMRI, Z-regression values for sensitivities and specificities were non-significant, suggesting that prediction success for structural or functional neuroimaging did not differ between studies (see **table 3)**.

**Table 3:** bivariate random-effect metaregression Z-scores for modality as covariate.

|  | Point estimate | Standard Error | 95% Lower | 95% Upper | z-value | p-Value |
| --- | --- | --- | --- | --- | --- | --- |
| Sens (lat) | 0.221 | 0.233 | -0.236 | 0.677 | 0.948 | 0.343 |
| Spec (lat) | 0.217 | 0.252 | -0.77 | 0.711 | 0.861 | 0.389 |

### 3.4 Quality assessment

Three studies included only late-life-depression, which reduces applicability in the general MDD population (see **supplementary figure 1** and **supplementary table 3**). In terms of flow and timing, drop-outs were a common issue, with 10 studies having a drop-out rate of 30 percent or higher, while 11 studies did not clarify drop-outs, possibly leading to attrition bias. Furthermore, two studies adapted the definition of response to create an even split in responders/non-responders, causing applicability concerns (42, 45). One study did not pre-specify the pharmacological intervention (47).

### 3.5 Heterogeneity and publication bias

The univariate forest plot of diagnostic performance (in ln OR) showed considerable overlap in confidence intervals between studies with different odds ratio’s, indicating that heterogeneity might be caused by sample variance (see **Figure 3**) (23). As described in the study description above, inter-study differences were present in population, modalities, intervention type, response/remission definition, feature selection and analysis technique. Deeks’ test showed study size and diagnostic odds ratio to be inversely related (p = 0.044) see **supplementary figure 2**), indicating that classification performance was lower in studies with larger samples. Inspection of the grey literature that was excluded due to missing information in order to construct a confusion matrix (all of which were conference/poster abstracts) showed that the grey literature had comparable mean sample sizes (n = 22, mean n = 56) and accuracies (ranging from 73%-95%) compared to the included studies. For an overview of grey literature results, see **supplementary table 4**.

**Figure 2:**
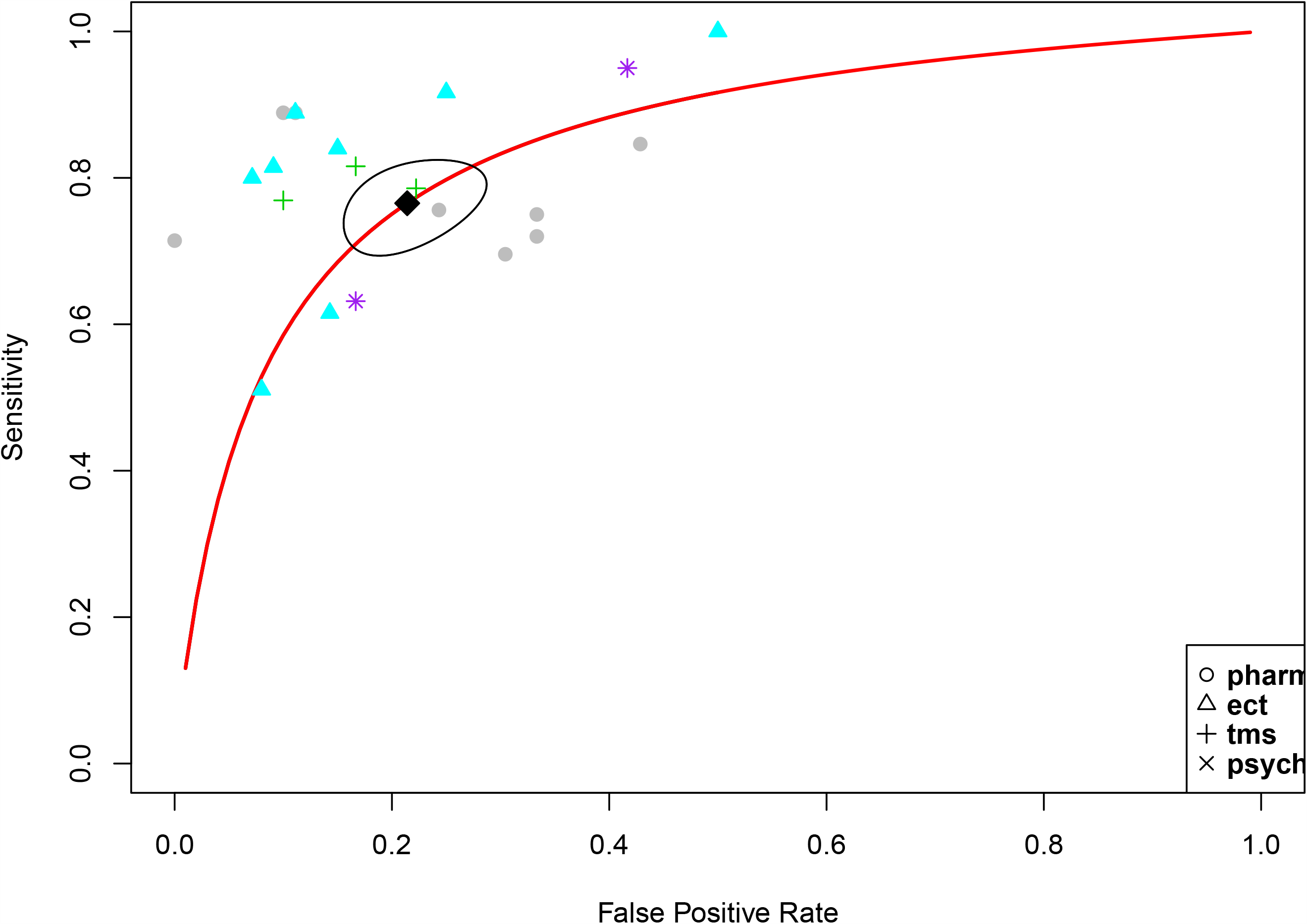
Meta-analysis of MRI-studies for prediction of antidepressant response, bivariate SROC. See PDF *Area under the bivariate summary receiver operating curve 0*.*84 (95% CI 0*.*81-0*.*87), sensitivity 77% (95% CI 71-82), specificity 79% (95% CI 73 - 84)*

**Figure 3:**
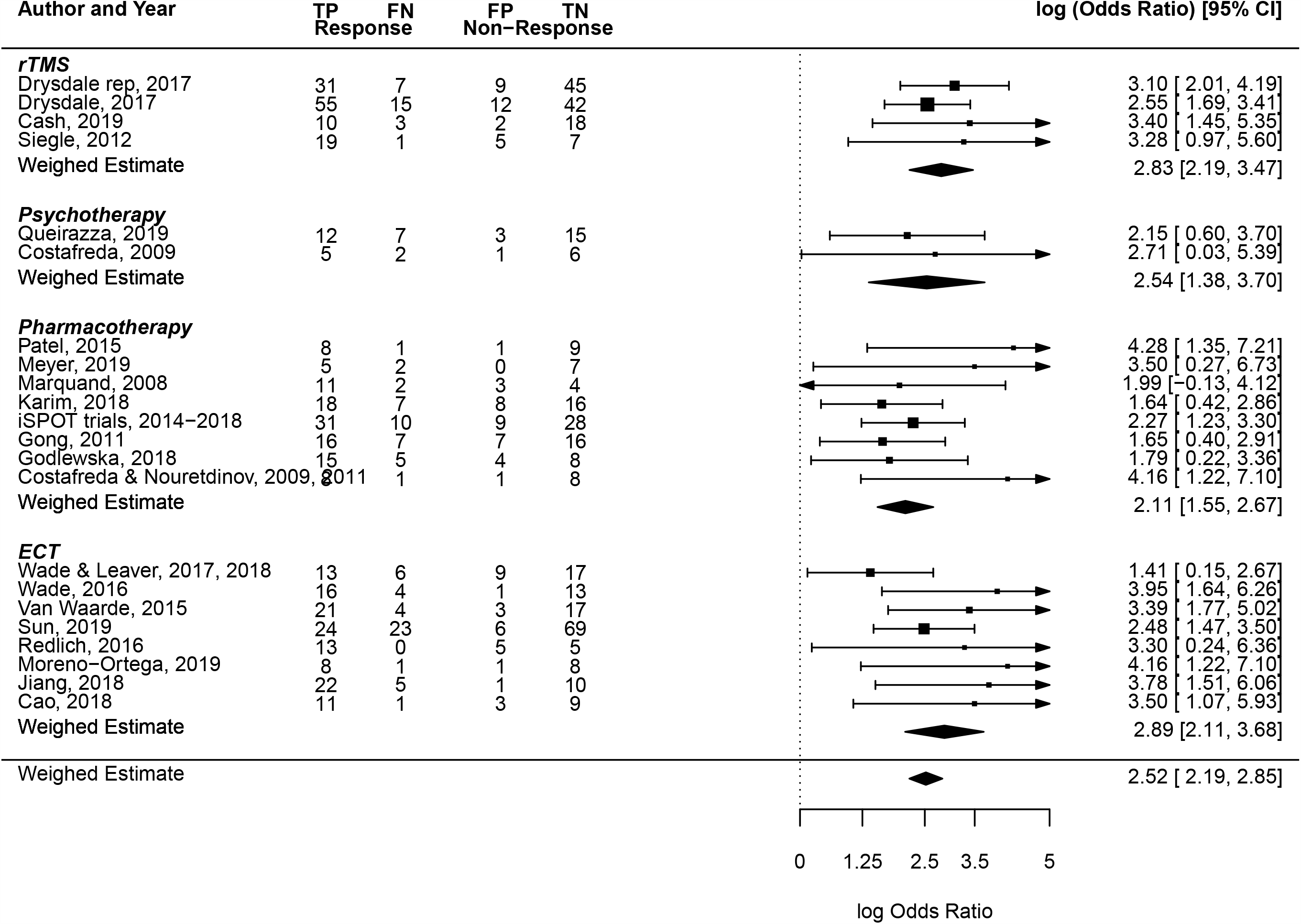
Univariate random-effect forest plot of ln diagnostic odds ratio’s. See PDF *Log(OR) = the natural logarithm (ln) of the odds ratio*.

## Discussion

Our results show that machine learning analysis of MRI data can predict antidepressive treatment success with an AUC of 0.84, 77% sensitivity, and 79% specificity (**figure 2**). Furthermore, we did not find a difference in classification performance between studies using pharmacotherapy and ECT. Although ECT showed somewhat higher sensitivity and specificity, confidence intervals largely overlapped between the two intervention types (**table 2**). In addition, classification performance of structural and functional MRI did not differ significantly (**table 3**).

To our knowledge, this is the first meta-analysis specifically examining MRI for predicting treatment effects in depression. The overall classification performance is comparable to the one reported by Lee et al., who found a general accuracy of 85% for neuro-imaging (defined as EEG, CT, PET or MRI) (54). Those results were however based on a total of 8 MRI-studies, whereas our search resulted in 21 individual samples for analysis. This is partly due to the time gap between studies, which underscores the rapid development in this research area. Our results show that MRI prediction studies perform somewhat better than EEG (AUC of 0.76) and comparable to accuracy of *diagnostic* classification studies with MRI that distinguish depressed patients and healthy controls (20, 55). In contrast to the review of EEG studies, we excluded studies that tested their model on the training set, which increased generalizability of our sample and avoided presenting inflated accuracy results.

Clinical practice would require different prediction approaches for a broad range of specific settings. It would be useful to have a single predictive test for therapy resistant patients, especially to guide decision making for invasive treatments such as ECT. For example, ECT is associated with cognitive side-effects that are preferably avoided in case the treatment is unsuccessful (56). In addition, ECT is only applied in 1-2% of patients with persistent or severe depression and a biomarker that indicates a high probability of success may reduce the hesitance of its use (57). However, for most treatments, a differential biomarker would be preferable, which would enable selecting the treatment with the highest chance of success. As of yet, no MRI-study has used such prospective prediction and subsequent treatment matching to guide decision making between two treatment options (for instance, between cognitive behavioral therapy and an SSRI). Furthermore, no studies have yet compared efficacy of prediction-guided treatments versus regular treatment based on patient-clinician preference. Thus, although the predictive performance of MRI biomarkers is certainly promising, the current study designs do not yet enable the translation of research findings to the clinic.

Generally, studies were of acceptable quality, although drop-out rates could cause concern in terms of reliability. Drop-out rates were not mentioned in 11 studies and for 10 studies, drop-out rates were larger than 30 percent without using an intention-to-diagnose approach. Not accounting for drop-outs, who might be less likely to respond to treatment, could inflate response/remission data and consequently alter sensitivity and specificity of the predictive test. Additionally, our results show between-study variety regarding the response criterion, which typically consisted of clinical response (≥50% symptom reduction) or symptom remission. Furthermore, although no objective investigation for clinical heterogeneity in prediction studies exists, our random-effect forest plot shows considerable overlap of confidence intervals with differing study results, implying the presence of sampling variation (**figure 3**) (22). Clinical variance between samples is an important obstacle in generalizability of any diagnostic or predictive marker, especially in psychiatric illnesses such as major depressive disorders, which is heterogeneous in both its clinical and neurophysiological manifestation (58, 59). Thus, inter-sample diversity of inclusion criteria and methodological design might hamper the realization of a reliable predictive biomarker.

In the current literature on diagnostic accuracy studies, the possibility of publication selection as a source of bias is still under debate (25, 60). However, our funnel plot (**figure 4**) shows the presence of a sample-size effect, with the *n* of a study being negatively correlated to classification performance, which could be attributable to publication bias (61). Another explanation of this significant correlation might be that large-scale studies with large samples are more likely to consist of heterogeneous patient groups, which in turn reduces prediction accuracy (62). As a further exploration of publication bias, our search also took into account grey literature, which indicated that publication (or positive result) bias was absent. In conclusion, quantitative testing could not distinguish between a real effect (due to accuracy reduction in large heterogeneous samples) or publication bias. Although the grey literature deems its presence less likely, we cannot exclude the presence of publication bias.

The following limitations warrant further discussion. First, we did not find modality differences, but studies conducting functional MRI research might have also attempted prediction with structural MRI which remained unpublished. Although we did contact authors for additional information, response was poor, so we were unable to rule out reporting bias for modality differences. Second, the number of studies predicting psychotherapy outcome was low, resulting in a blind spot for one of the most commonly deployed treatment modalities in MDD (63). Third, cross-validation in small samples results in large variation of the estimated accuracy and as indicated above, accuracy reduces with larger sample heterogeneity (62, 64). Since the mean sample size of our studies was 44 (with a median *n* of 33), the reported results may be optimistic. Furthermore, characteristics of the test set during cross-validation will approximate the characteristics of the training set more than when tested in the general population, due to selection bias (65). Only two included studies replicated their training data in an independent cohort, and one included study used an out-of-sample cohort to further test their cross-validated results, leaving the question open to which extend the majority of results can be generalized to new patients (30, 36, 50).

In order to optimize patient care, reduce treatment resistance and shorten duration of illness, developing models that predict treatment success on individual-patient level is an urgent task. In a 2012 consensus report on diagnostic imaging markers in psychiatry, the American Psychiatric Association research council proposed 80% sensitivity and specificity as prerequisite for the clinical application of a biomarker (66). Furthermore, biomarkers should be ideally be reliable, reproducible, non-invasive, simple to perform, and inexpensive. The results for an ECT biomarker fulfilled the 80% criterion, but the results for a medication biomarker fell short. But following these terms, primarily reproducibility has not yet been sufficiently well established with small sample-sizes and external validation in only a minority of studies. This precludes recommending MRI for treatment response prediction in clinical practice at this point.

Future multicenter studies with large patient samples that represent clinical heterogeneity are required to warrant MRI biomarker generalizability (67). However, one might question whether excellent generalizability is a goal which should be aimed for: if each clinical site were to develop its own, locally reliable and replicable biomarker that incorporates the local hardware, patient, and treatment variability, the predictive accuracy is expected to be higher than when all potential sources of heterogeneity are accounted for (62, 68). Standard machine learning analysis would, then, mean a departure from the traditional universalist paradigm in diagnostics, and instead initiate a shift to a paradigm of localization: heterogeneous yet locally applicable classification models. This will enable to retrain predictive models to obtain even better performance with more data after biomarker deployment. And this may enable to take advantage rather than disadvantage from (inevitable) hardware upgrades, such as higher signal-to-noise for new generations of MR scanners and coils.

In conclusion, prediction of treatment success using machine learning analysis of MRI data holds promise but has not transcended the research status and should not yet be implemented into clinical practice. Once it overcomes the aforementioned hurdles, MRI may become a clinical decision support tool aimed to reduce unsuccessful treatments, and improve treatment efficacy and efficiency.

## Data Availability

All data available are included in our manuscript.

## Supplementary material

### Supplementary methods

#### Search Strategy

Our search strategy included terms regarding the population, the diagnostic predictor and the study results, in a measure of accuracy. Studies were included up to January 2020. We used no language or date and we excluded animal/non-human studies. The search was conducted by a clinical librarian and search specialist (JD), in order to ensure a high degree of thoroughness.

We searched the following electronic databases: EMBASE, Medline and PsycInfo and Web of Science. Our Medline search was constructed as follows:

((depression/ or postnatal depression/ or major depression/ or treatment resistant depression/) OR (mdd or major depressi* or unipolar or postnatal depressi* or post natal depressi* or postpartum depressi* or post partum depressi* or refractory depression or (depressi* adj3 resistan*) or late life depressi*).ab,kw,ti.)) AND

((nuclear magnetic resonance imaging/ or diffusion tensor imaging/ or exp neuroimaging/ or functional magnetic resonance imaging/) OR ((magnetic resonance or mr imaging or mri or fmri or dti or diffusion tensor or tensor imaging or structural neuroimaging or functional neuroimaging or structural neuranatomy or functional connectivity).ab,kw,ti.)) AND

((“sensitivity and specificity” / or predictive value/ or exp diagnostic error/ or diagnostic accuracy/) OR ((“sensitivity and specificity” or npv or ppv or roc or predict* or prognos* or accuracy).ab,kw,ti.))

Additionally, we checked every included article for relevant references. As of yet no central registration database exists for unpublished diagnostic studies, however we searched in the WHO International Clinical Trial Registry Platforms search portal for registered and unpublished studies. Furthermore, we looked for ‘grey’ literature such as (poster) abstracts and conference articles through conference websites (SOBP, ISAD, APAAM, ACNP, ECNP, HBP, WCP, Molecular Psychiatry, ADAA, going back to 2009) and from other relevant sources.

Our inclusion criteria, as well as our meta-analytic methods of procedure were pre-registered in the PROSPERO register of systematic reviews (registration ID CRD42019137497).

#### Inclusion and exclusion

##### Inclusion criteria

- Adults (18 years or older) diagnosed with Major Depressive Disorder, as diagnosed using the DSM III, IV or 5 criteria. We included the entire MDD-population, including all severity subtypes. Therapy resistance status was allowed to range from naive to resistant. We included both in-and outpatients.
- Magnetic Resonance Imaging (structural MRI, task-based functional MRI, resting state functional MRI, diffusion tensor imaging) before the start of antidepressive treatment (all-compassing, ranging from psychotherapy to pharmaceutical treatment or electroconvulsive therapy, as included in the National Institute for Health and Care Excellence Guidelines for antidepressive treatment) [3]. To ensure that data used for treatment prediction correspond with the situation at treatment-baseline, MRI had to be performed within a month before treatment commencement date.
- Accuracy of prediction had to be evaluated by comparing the predicted outcome, to a validated disease-severity questionnaire or semi-structures interview *after* treatment. Such questionnaires/interviews included: the Hamilton Depression Rating Scale −17, the Beck Depression Inventory, the Montgomery Asberg Depression Rating Scale or the (Quick) Inventory of Depressive Symptomatology, specified as either self-rated or clinician rated. We considered the validity of all these rating scales to be equal. Furthermore, we included studies that measure symptom severity/response status within 12 weeks after treatment commencement. We chose this cut-off to allow pharmacotherapeutic dose escalation corresponding with routine clinical practice, and to allow time for psychotherapeutic therapies to take effect.
- We included studies that set out to predict, on the level of the individual patient with a depressive episode, response or remission to therapy. We did allow for any therapy that is part of official treatment guidelines, since our main aim is to investigate the general possibility of MRI to predict therapeutic response. We allowed for psychiatric co-medication such as benzodiazepines, since these are administered routinely in daily clinical practice.
- We included any study with a pre-specified definition of response/remission.
- We allowed studies to have defined prediction as sensitivity/specificity, positive or negative likelihood ratio, positive or negative predictive value, a measure of overall accuracy, the area under the ROC-curve, a Youden’s index, diagnostic odds ratio or any other measure that illustrates accuracy and/or may be used to compute a confusion matrix.

##### Exclusion criteria

- We excluded studies with patients younger than 18 years old, or patients with a bipolar disorder. We chose to exclude patients with a depressive episode in context of a bipolar disorder, since there is evidence that bipolar depression exhibits characteristics which significantly differ from unipolar depression, both clinically and neurobiologically [1, 2]. If an article included a mixed sample of bi- and unipolar depressant patients, we excluded the study if more than two-thirds of patients suffered from bipolar disorder
- If studies used feature selection based on in-sample data, we excluded them if they did not validate their prediction outcome either internally (e.g. through cross-validation) or externally (through independent set validation). We chose this approach since accuracy measures based on data that are included in model training are positively biased and have unknown (and presumably low) levels of generalizability, deeming them irrelevant for making a substantiated recommendation, as was the goal of our review. Studies that did not validate their data, but did use a-priori defined methods and features for analysis, were included.

#### Data extraction

Two authors (SC and BW) extracted, if provided, from included studies the following data: number of participants, gender distributions (male/female), age, mean severity pre-intervention, number of prior episodes, duration of current (index) depressed episode, whether or not patients had a history of failed antidepressant treatment, if patients used current psychiatric medication (and if so, which ones), specific exclusion criteria. Furthermore, we specified treatment, dosage/frequency, duration of treatment, defined endpoint and response or remission rates. For the diagnostic tests, we specified modality, region of interest, machine-learning algorithm, cross-validation analysis. As results, we extracted sensitivity, specificity and the confusion matrix (true prositive, false negative, false positive, true negative). For the full result table, please refer to **supplementary table 1**.

#### Quality analysis

For risk of bias in patient selection we asked if a consecutive or random sample of patients enrolled and if the study used proper exclusion criteria. For the index test (i.e. the MRI biomarker) we asked if multiple comparison testing was used for feature selection. For the reference standard (i.e. the questionnaire or rating scale) we asked if the reference standard was validated for MDD. For flow and timing, we considered an appropriate interval between MRI and questionnaire (i.e. within 12 weeks), and whether all patients did indeed receive the same rating scale. Furthermore, we considered which percentage of patients who underwent an initial predictive MRI, did indeed finish the treatment protocol. If there was a drop-out rate of more than 30%, we considered this as a high risk of bias. We added a section intervention, in which we took into account whether the intervention (kind, dosage, duration) was pre-specified at baseline.

Studies that selected patients who did not solely include MDD patients (i.e. also included a small portion of depressed bipolar patients) or who included a specific subsection of MDD patients, such as late-life depression, were registered as having a high applicability concern. Furthermore, if the index test (i.e. the MRI) were to be combined with clinical data other than age, disease severity and sex, we considered concerns to be ‘high’. Cut-offs of response/remission the reference standard (i.e. rating scale/questionnaire) were preferable pre-specified and if not, applicability concerns were high. If for the intervention, frequency, dosage and time did not match clinical standards, applicability concerns were high.

#### Meta-analytic procedure

We pooled studies using a bivariate random effect model according to Reitsma, as suggested in the Cochrane handbook for diagnostic tests of accuracy studies [34, 36]. Main outcomes were the overall area under the sROC-curve and sensitivity/specificity, as well as sens/spec of intervention subset (pharmacological treatment, specified in appropriate considering amount of studies, electroconvulsive therapy, psychotherapy, transcranial magnetic stimulation and any other intervention with a 5 or more studies). A cause of heterogeneity in diagnostic or predictive tests is the threshold effect; in prediction models, studies use varying cut-offs for classifying a patient as responder or remitter. This threshold effect causes an inverse relation between sensitivity and specificity. Most neuroimaging studies establish empirically which threshold should will used for prediction, based on which produces the highest overall accuracy. The bivariate sROC curve is specifically suitable for this kind of heterogeneity since its visual properties take the correlative relation between sensitivity and specificity into account [35]. For computing confidence intervals for the area under the ROC-curve, we used the method of Hanley and McNeil (1982).

To detect sample size effect and possible publication bias, we plotted a funnel plot based of 1/√ (effective sample size) as a function of the natural logarithm of the diagnostic odds ratio [43]. Known as the Deeks’ test, this function is the recommended test for sample size effect [44]. More common forms of formalizations of publication bias, such as the Egger’s or Begg’s test are not recommended for this review, since their sensitivity for diagnostic accuracy studies is generally poor [36, 43] [35].

#### QUADAS quality assessment

<u>Risk of bias</u>

1. Patient selection
  - Was a consecutive or random sample of patients enrolled?
  - Did the study avoid inappropriate exclusions?
2. Index test
  - Were the index test results interpreted without knowledge of the results of the reference standard?
  - If a threshold was used, was it pre-specified?
3. Reference standard
  - Is the reference standard likely to correctly classify the target condition?
  - Were the reference standard results interpreted without knowledge of the results of the index test?
4. Flow/timing
  - Was there an appropriate interval between index test and reference standards?
  - Did all patients receive a reference standard?
  - Did patients receive the same reference standard?
  - Were all patients included in the analysis?
5. Intervention

- Was the intervention (kind, dosage, duration) pre-specified?

<u>Applicability</u>

1. Risk of bias
  - Is there concern that the included patients do not match the review question?
2. Index test
  - Is there concern that the index test, its conduct, or interpretation differ from the review question?
3. Reference standard

- Is there concern that the target condition as defined by the reference standard does not match the review question?

## Supplementary discussion

### Discussion of quality assessment

Not one study used a consecutive patient enrollment, causing concern for selection bias and thus low applicability. A problem might arise specifically when researchers choose not to include certain patients for their study, on the basis of reasons outside of a-priori protocol. Such reasons might include expectations about whether patients will be reliable in finishing the study, co-morbid substance abuse or predominantly ‘psychosocial’ events leading up to depressive disorder. Consecutive enrollment is not an issue generally discussed within diagnostic studies, but since our included studies are at the same time intervention studies, these concerns might be especially relevant.

Furthermore, three studies included only late-life-depression, reducing applicability in the general MDD population, as late-life-depression, although being symptomatically similar with MDD in a lot of aspects, is thought to co-occur more often with medical disorders such as vascular illnesses and neurocognitive disorders.

In the terms of flow and timing, drop-out-rates, as discussed in the main body of text, cause issues around attrition bias. All studies used multiple correction control in feature selection for the index test, through the process of cross-validation, as was one of the inclusion criteria. One study did not pre-specify the pharmacological intervention (Patel et al.). Two studies adapted their definition of response to create an even split in responders/non-responders, causing applicability concerns since one would have to know exactly which predefined outcome to predict for response prediction to be clinically relevant (Meyer et al., 2019; Leaver et al., 2018).

### Non-validated studies

Remarkably, we did exclude as much as eleven articles that did not use a form of validation after they used their training data for response prediction. In most of these cases, dichotomous response prediction was a secondary, or post-hoc, analysis, and some authors do note that these results might not be generalizable or should be interpreted with caution. However, all authors did report their results in the abstract, clouding research on this topic. Furthermore, for a clinician, critically evaluating the absence of cross-validation or independent test set validation is infeasible which could lead to overestimation of prediction success. Therefore, we would advise against publishing non-validated prediction results if these studies use feature selection in the training sample.

### Feature selection and analysis

We did not discuss different machine-learning methods or feature-selection techniques in depth, since these issues go beyond the scope of our research question. However, summary of our results (see **supplementary table 2**) indicates that no single region-of-interest, network-analysis or functional task seems to stand out in prediction success, although with our data we could not quantitatively substantiate this finding. For instance, one found an important predictive role for the subcallosal cingulate gyrus, while another, after whole-brain data mining, did not include this brain part in their final predictive model [57, 59]. Thus, cerebral processes which influence treatment success seem to be overdetermined and prediction might be made in a multitude of ways

## Supplementary tables

**Supplementary table 2:** Imaging and regions of interest. PCA = principal component analysis, ROI = region of interest, SVC = small-volume correction, FC = functional connectivity, DTM = diffusion tensor model, ICA = individual component analysis, CCA = canonical correlation analysis, GLM = general lenear modeling, WB = whole-brain, VBM = voxel-based morphometry, FG = frontal gyrus, TG = temoral gyrus, CgC = cingulate gyrus, sgACC = subgenual anterior cingulate cortex, pgACC = pregenual anterior cingulate cortex, iOFC = inferior orbitofrontal cortex, Cd = caudate, PCL = paracentral lobule, TPJ = tempoparietal junction, amPFC = anteriomedial prefrontal cortex, dl = dorsolateral, dDMN = dorsal default mode network, aSN = anterior salience network, iTC = enferior temporal cortex, OcC = occipital cortex, MFGL = mediofrontal gyrus, FTC = frontotemporal cortex, CA = hippocampus, ST = stria terminalis, CBPM = connection-based predictive modelling

| Modality | Study + year | Analysis approach | ROI's / top regions | Task |
| --- | --- | --- | --- | --- |
| **tbfMRI** |  |  |  |  |
|  | Marquand, 2008 | PCA | FG, TG, CgC, midbrain, cerebellum, precuneus | 3-back verbal memory |
|  | Siegle, 2012 | ROI, hypothesis-driven | sgACC | personal relevance rating |
|  | Williams, 2015 | ROI, hypothesis-driven | amygdala | facial emotion paradigm |
|  | Goldstein-Piekarski, 2016 | ROI, hypothesis-driven | amygdala | facial emotion paradigm |
|  | Godlewska, 2018 | ROI, hypothesis-driven, SVC | pgACC | facial expression, masked |
|  | Karim, 2018 | PCA | left iOFC, hippocampus., bilateral FG, left Cd, right PCL | facial expression / shapes |
|  | Kraus, 2018 | Gaussian kernel | right TPJ | pain anticipation |
|  | Meyer, 2019 | Context-independent FC | amPFC, dlPFC, , pCgC | n-back working memory |
|  | Queirazza, 2019 | Gaussian kernel | right amygdala, right striatum | Reversal-learning |
| **rsfMRI** |  |  |  |  |
|  | Patel, 2015* | ROI hypothesis-driven, DTM computation, PCA | dDMN, aSN |  |
|  | Van Waarde, 2015 | ICA | brainstem, CB, dlPFC, dmPFC, iTC, OFC |  |
|  | Drysdale, 2015 | CCA | 25 regions of interest |  |
|  | Leaver, 2017 | ICA | 23 regions of interest |  |
|  | Wade, 2017 | Automated volumetric parcellation (FreeSurfer) | CA, iTC |  |
|  | Goldstein-Piekarski, 2018 | ROI, hypothesis-driven, GLM | amygdala |  |
|  | Moreno-Ortega, 2019 | Multimodal parcellation | dlPFC |  |
|  | Sun, 2019 | Functional connectome, CBPM | Top features: Thalamus, temporal/hippocampal/frontal gyri |  |
| **sMRI** |  |  |  |  |
|  | Costafreda, 2009 | Gaussian kernel | CgC, Occ, MFG |  |
|  | Nouretdinov, 2011 | Gaussian kernel | CgC |  |
|  | Gong, 2011 | WB, high-dimensional normalization protocol | Grey/white matter; FTC, Occ,, putamen |  |
|  | Redlich, 2016 | WB, high-dimensional normalization protocol | Grey-matter integrity |  |
|  | Wade, 2016 | VBM, GLM | CA, iTC |  |
|  | Cao, 2018 | Automated volumetric parcellation (FreeSurfer) | CA |  |
|  | Jiang, 2018 | VBM, unified segmentation | CA, left FTG, left LG, left precuneus |  |
| **DTI** |  |  |  |  |
|  | Korgaonkar, 2014 | DTM | CgC, ST |  |
|  | Grieve, 2016 | ROI, hypothesis-driven, DTM | CgC, ST |  |

**Supplementary table 3:** QUADAS quality assessment. ? = unknown (not mentioned in article), - = high risk of bias or low applicability, + = low risk of bias or high applicability. MMSE = mini mental state examination, LLD = late-life depression, non-rem = non-remission

| Study | Risk of bias |  |  |  |  | Applicability concerns |  |  |  |
| --- | --- | --- | --- | --- | --- | --- | --- | --- | --- |
|  | Patient selection | Index test | Reference standard | Flow and Timing | Intervention | Patient selection | Index test | Reference standard | Intervention |
| Marquand, 2008 | ? | + | + | - | ? | + | + | + | ? |
| Costafreda, 2009 | ? | + | + | ? | + | + | + | + | + |
| Costafreda 2, 2009 | ? | + | + | ? | + | + | + | + | + |
| Nouretdinov, 2011 | ? | + | + | ? | + | + | + | + | + |
| Gong, 2011 | ? | + | + | - | + | + | - | + | + |
| Siegle, 2012 | ? | + | + | - | + | + | - | + | + |
| Patel, 2015 | ? | + | + | ? | - | - | - (MMSE) | + | + |
| Van Waarde, 2015 | ? | + | + | + | + | + | + | + | + |
| Redlich, 2016 | ? | + | + | + | + | + | + | + | + |
| Wade, 2016 | ? | + | + | ? | + | ? | + | + | + |
| Drysdale, 2016 | ? | + | + | + | + | + | + | + | + |
| Wade, 2017 | ? | + | + | ? | + | ? | + | + | + |
| Leaver, 2018 | ? | - | - | ? | + | ? | + | - | + |
| Cao, 2018 | - | + | + | ? | + | - | + | + | + |
| Godlewska, 2018 | ? | + | + | - | + | + | + | + | + |
| Jiang, 2018 | ? | + | + | + | + | - (LLD) | + | + | + |
| Karim, 2018 | ? | + | + | - | + | - (LLD) | + | + | + |
| Cash, 2019 | ? | + | + | + | + | + | + | + | + |
| Meyer, 2019 | ? | - | - | + | + | + | + | - | + |
| Moreno-Ortega, 2019 | ? | + | + | ? | ? | - | + | + | ? |
| Queiraza, 2019 | ? | + | + | ? | + | + | + | + | + |
| Sun, 2019 | ? | + | + | ? | + | ? | + | + | + |
| Korgaonkar, 2014 | ? | + | + | - | + | + | + | + | + |
| Williams, 2015 | ? | + | + | - | + | + | + | + | + |
| Goldstein-Piekarski, 2016 | ? | + | + | - | + | + | + | + | + |
| Grieve, 2018 | ? | + | + | - | + | + | - (non-rem) | + | + |
| Goldstein-Piekarski, 2018 | ? | + | + | - | + | + | + | + | + |

**Supplementary table 4:** Overview of grey literature. N = number of participants, AUC = area-under-the curve, rsfMRI = resting-state functional MRI, rTMS = repetitive transcranial magnetic stimulation, tbfMRI = task-based fMRI, SSRI = selective-serotonin reuptake inhibitor, CBT = cognitive behavioral therapy

| Name + year | N | Accuracy | AUC | Sensitivity | specificity | Modality | intervention |
| --- | --- | --- | --- | --- | --- | --- | --- |
| Cash, 2019 | 47 | 85-95% |  | - | - | rsfMRI | rTMS |
| Etkin, 2013 | 102 | - |  | - | - | tbfMRI | SSRI |
| Fitzgerald, 2019 | 120 | >85% |  | - | - | fMRI | rTMS |
| Geraci, 2014 | - | 83, 90% |  | - | - | fMRI | rTMS |
| Godlewska, 2017 | 32 | 75% |  | - | - | fMRI | SSRI |
| Hou, 2016 | 82 | - | 0.69, 0.71 | - | - | fMRI | - |
| Karim, 2018 | - |  |  | 80 | 63 | fMRI | SSRI |
| Klobl, 2019 | 35 | - | 0.73 | - | - | fMRI | SSRI |
| Klumpp, 2019 | 20 | 80% | - | - | - | tbfMRI | CBT |
| Korgaonkar | 157 | 85% |  | - | - | fMRI | SSRI |
| Kozel, 2010 | 13 | 85% |  | - | - | fMRI | rTMS |
| Langenecker, 2014 | 24 | - | - | - | - | fMRI | SSRI |
| Long, 2019 | 59 | - | - | - | - | fMRI | rTMS |
| Miller, 2018 | 31 |  |  | - | - |  |  |
| Narr, 2016 | 22 | 76-77% | 0.75-0.80 | - | - | fMRI | ECT |
| Nguyen, 2019 | 37 |  | 0.71 |  |  | fMRI | bupropion |
| Schultz, 2018 | 21 | 88.95% | - | - | - | tbfMRI | - |
| Siegle | 53 | - | - | - | - | fMRI | SSRI |
| Vila-Rodriguez, 2018 | 62 | 76%-84% | 0.75-0.87 | - | - | fMRI | rTMS |
| Wade, 2015 | - | - | - | - | - | fMRI | ECT |
| Webb, 2018 | 35 | - | - | - | - | fMRI | CBT |
| Williams, 2013 | 101 | - | - | - | - | fMRI | SSRI |

**Supplementary figure:**
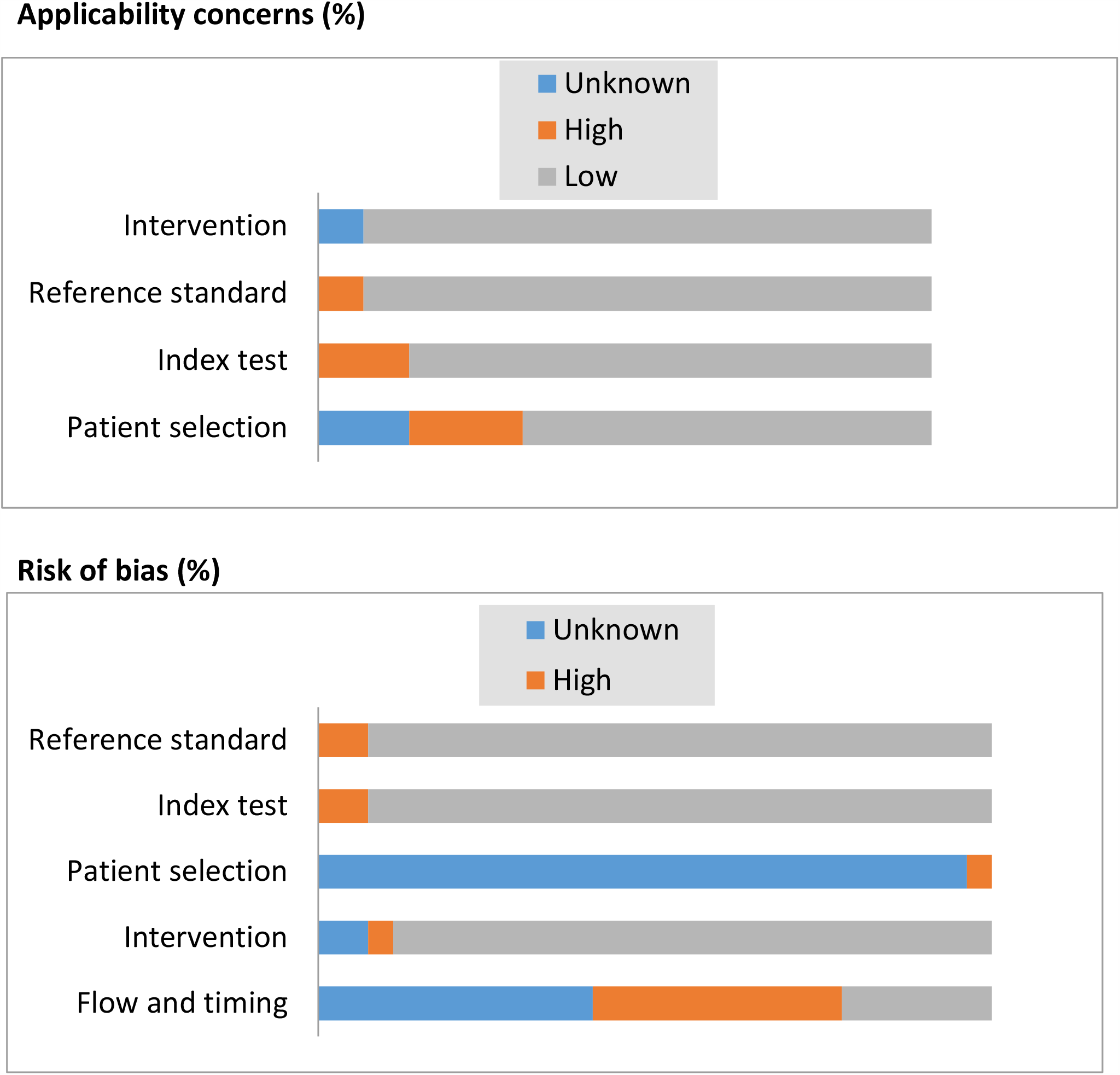
QUADAS-2 quality assessment.

**Supplementary figure 2:**
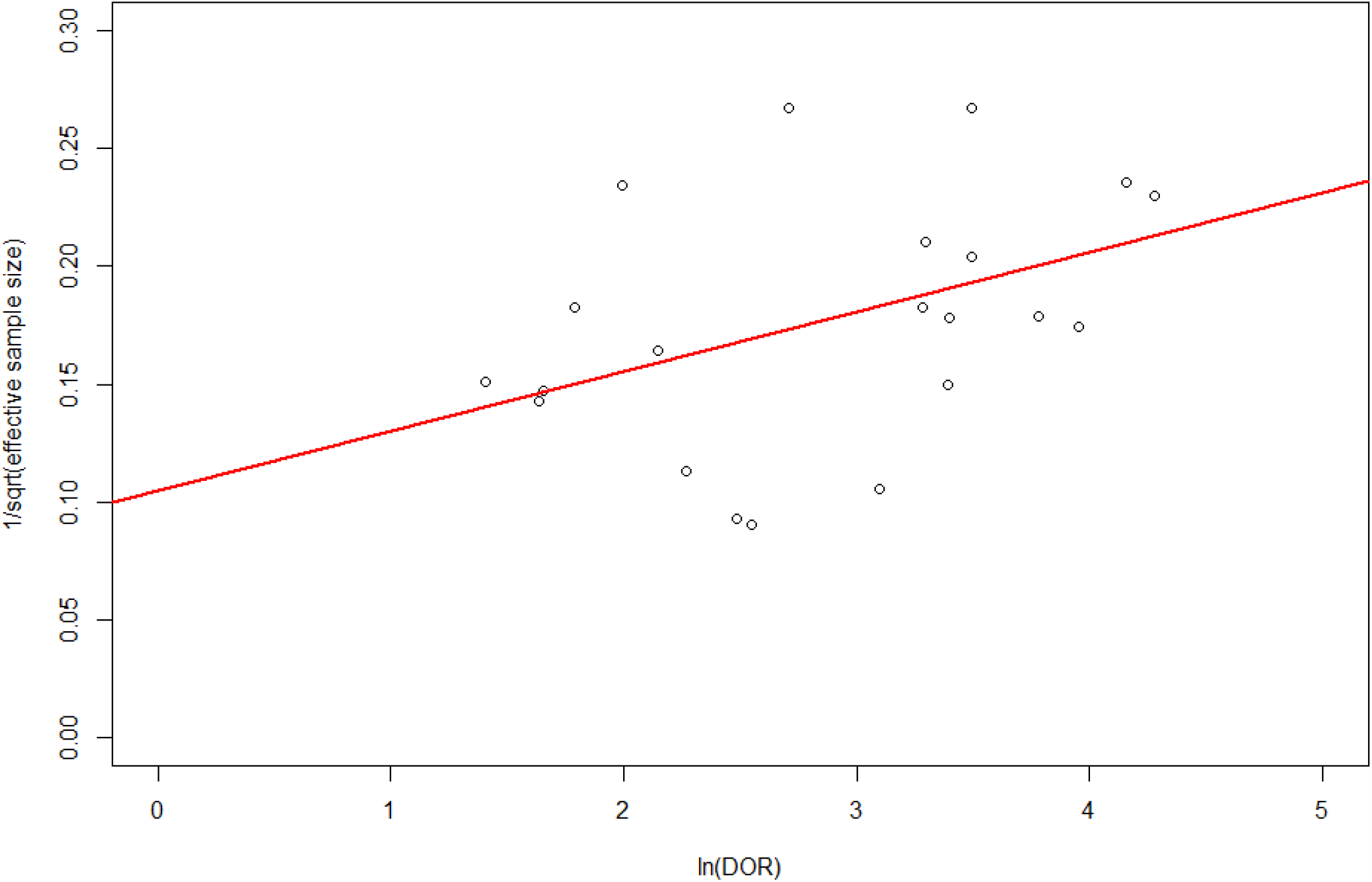
Deeks’ test, or sample size effect: 1/ √Effective Sample Size = (4xRxNR)/(R+NR) as a function of the ln Diagnostic Odds Ratio of each study. Regression equation: y = 0.11 + 0.025x, **p = 0**.**044** *The positive correlation between ln(DOR) and 1/√ESS indicates an negative correlation between DOR and ESS. The larger the sample size becomes, the smaller is the diagnostic accuracy. (R = responder, NR = non-responder)*.

**Supplementary table 1:**
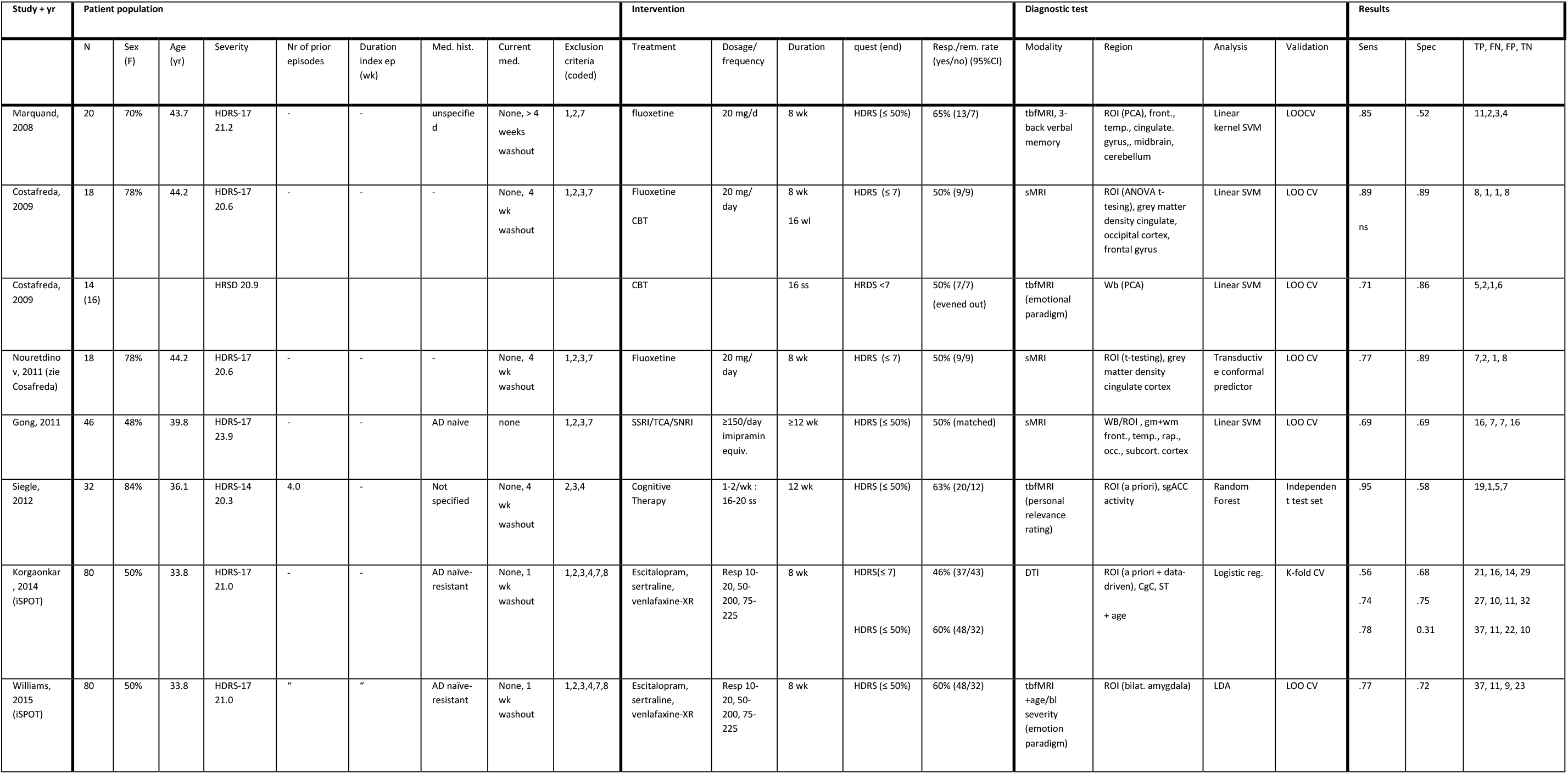
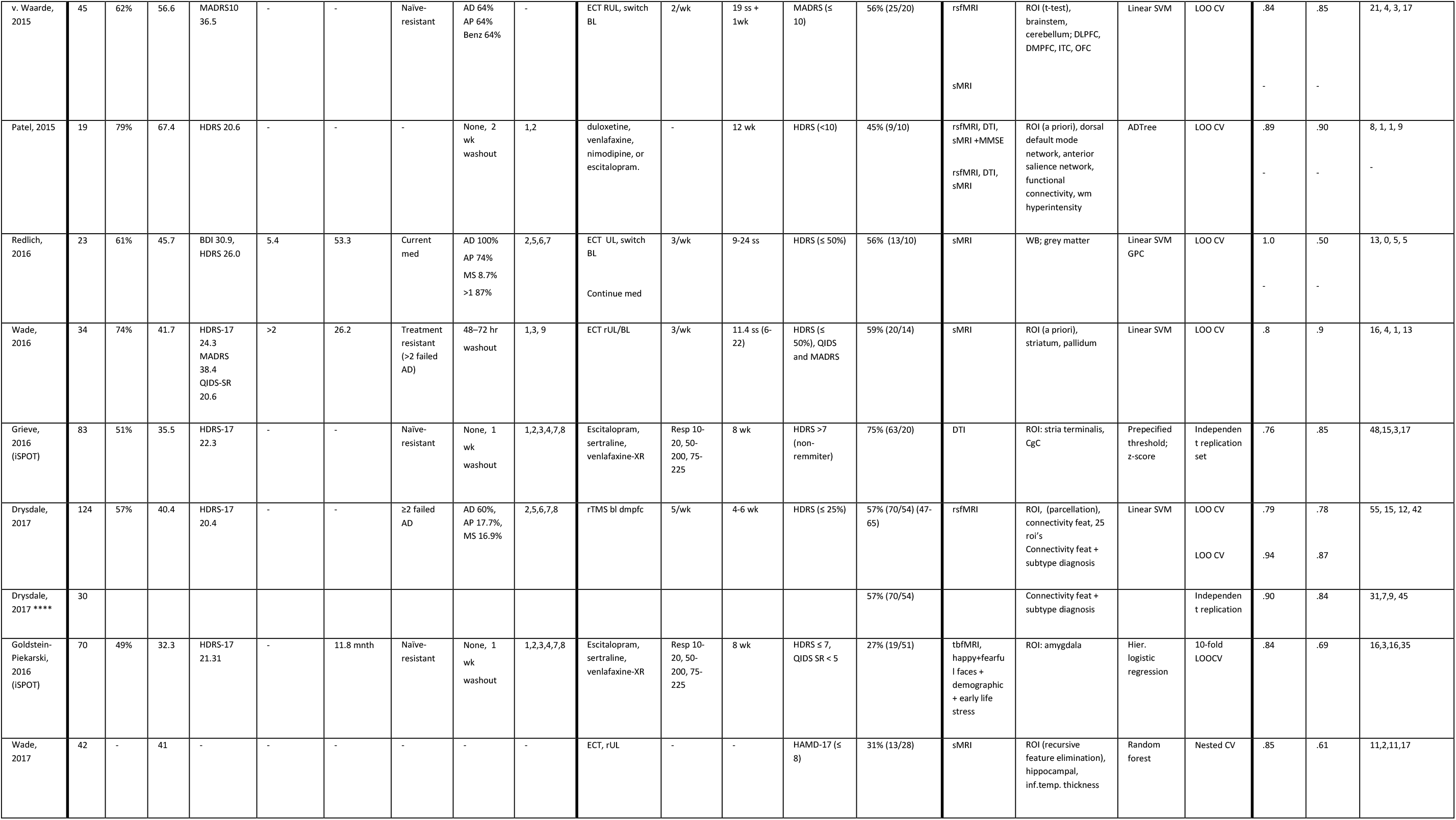
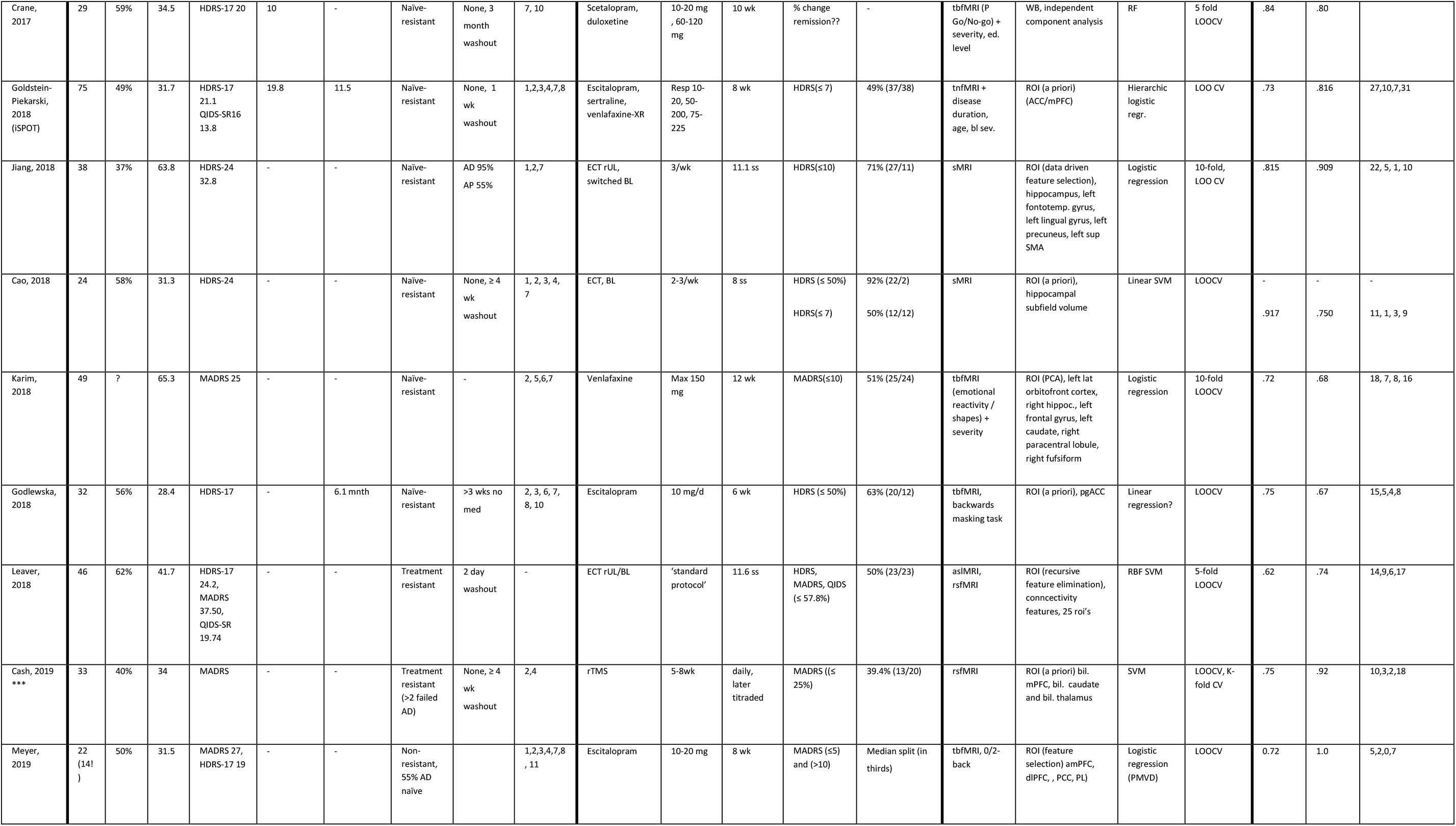
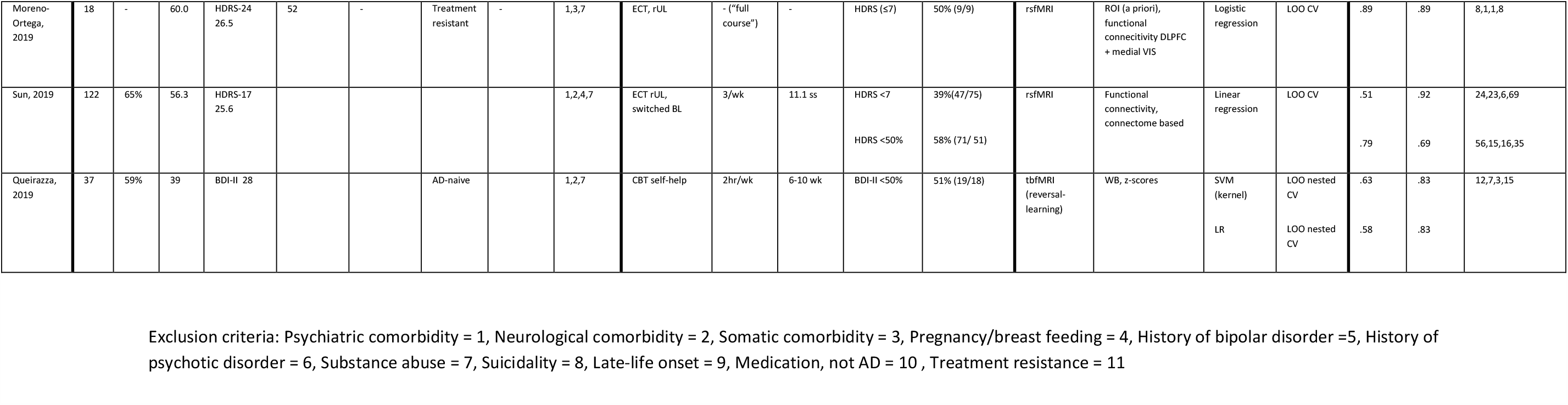
all-encompassing study summary.

## Notes

### Competing Interest Statement

The authors have declared no competing interest.

### Funding Statement

This research did not receive any specific grant from funding agencies in the public, commercial, or not-for-profit sectors.

### Author Declarations

Our research did not use primary human data since it was a meta-analysis.

